# Stem Cell Transplantation Patients and Situational Meaning: A Prospective and Longitudinal View

**DOI:** 10.1101/19001529

**Authors:** Jennifer Blosser, Roy Sabo, Kathryn Candler, Karen Mullin, Amir Toor, Denna Chaber

## Abstract

**Background:** Though there is thorough examination of psychosocial issues in SCT, there are few longitudinal studies examining the meaning SCT patients attribute to their treatment.

**Objective:** The object of this study was to examine change in situational appraisal over time, and to explore potential modifiers of that change.

**Methods:** A prospective, longitudinal study of 146 autologous and allogeneic stem cell transplant (SCT) patients at Virginia Commonwealth University (VCU) was conducted to measure situational appraisal over time as per the meaning making model of Park and Folkman (1997) utilizing the Illness Perception Questionnaire-Revised (IPQ-R; Moss-Morris et al., 2002). Participants were administered the instrument prior to transplant, at one, three and six months, and at one-year post-transplant.

**Results:** Change over time was seen in different diagnoses, donor types, reduced intensity and standard pre-transplant preparative regimens, and between the two major ethnic groups (Caucasian and African American) studied. Many of the patient subgroups had statistically significant findings in measures of illness attribution.

**Conclusion:** The appraisals SCT patients made of their situation in treatment revealed a complex process of appraisal affected by illness, treatment and patient characteristics including disease type, donor type, race, and pre-transplant regimen.

## INTRODUCTION

Patients undergoing stem cell transplant (SCT) are surviving and living longer with varying levels of transplant morbidity (Stronach, 2017; Leukemia & Lymphoma Society, 2017; American Cancer Society, 2017; Clauser, Gayer, Murphy, Majhaul & Baker, 2015; Patient-Centered Outcomes Research Institute, 2017). The acute transplant process presents numerous medical complications and significant toxicity. Increase in survivorship is a result of improvements in medical science (Stronach, 2017; Leukemia & Lymphoma Society, 2017; American Cancer Society, 2017). Patients experience a variety of adverse psychological reactions during this treatment, and face existential crises, including fear of death, loss of control, isolation, increases in dependency, and disabling physical symptoms. The transplant process is rigorous and invasive, creating trauma that can disrupt a patient’s sense of meaning as defined by the meaning making model of Park and Folkman (1997), who defined meaning making as an intrinsic emotional/psychological processing or an extrinsic attempt to make sense of life events (see the meaning making model chart in supplementary material).

Most of the literature on the emotional experience of SCT patients focuses on quality of life after transplant, survival factors, social support, psychological reactions, candidacy assessment, as these are primarily based in bio-psychosocial theory and explore various aspects and combinations of these factors. The authors were able to find a few articles that closely examined the existential crises associated with transplant (Beanlands, Lipton & McKay, 2003; Cohen& Ley, 2000; Jones & Chapman, 2000; Morstyn, 2009; Quinn, 2003; Xuereb & Dunlop, 2003). Moore and Goldner-Vukov (2009) state that, “The bio-psychosocial model often fails to sufficiently validate the existential suffering of patients.” The experience of existential suffering and the process of meaning making have been united in concept and process in the experience of cancer patients (Breitbart, 2017). However, there are few examples of longitudinal studies examining situational illness appraisals over time. Situational appraisal is defined as an internal cognitive assessment process of a personal - environmental transaction in which one’s sense of concordance or discordance with beliefs and abilities can generate emotion and affect coping. (Park & Folkman, 1997). The appraisal made of a situation as discrepant with global meaning can cause distress and leads to higher risks of negative emotional outcomes (Park & Folkman, 1997). The unresolved distress can lead to altered coping and adjustment; conversely, successful resolution of distress facilitates post traumatic growth and the resultant successful adjustment (Park, Chmielewski & Blank, 2010). The rigor of stem cell transplantation causes patients to initiate meaning making attempts due to the disruption and distress it creates.

This study attempted to increase understanding of a theoretically complex process of appraisal and adjustment by focusing on the patient experience. The meaning making model posits that individuals have a sense of global meaning, defined as beliefs/goals about a just world; this meaning is shattered by traumatic situations, leading to distress. Individuals engage in a process to rectify this and reduce distress, making intrinsic and extrinsic attempts to make sense of the disparity resulting either in continued distress or resolution. Successful resolution is seen in assimilating the new information into pre-existing schemata or accommodating – essentially revising one’s global meaning. Distress has been studied from the perspective of meaning making as an attempt to understand significant life stressors (Park, 2010). The meaning making model was the theoretical basis for examining a sample of SCT patients at VCU in a prospective, longitudinal behavioral health study examining changes in situational appraisal over time, and also to explore potential modifiers of that change.

### TRANSPLANT AND EMOTIONAL WELL-BEING

Stem cell transplant is the process by which a person’s bone marrow is either partially or totally ablated by combinations of chemotherapy, radiotherapy, and immune suppressive medicine and the resulting aplasia managed by an infusion of their own previously harvested marrow/ stem cells (autologous transplant) or a donor’s marrow / stem cells (allogeneic transplant). Hematological cancers are the primary illnesses treated in this fashion. Following transplantation, patients make frequent visits to the transplant center for close medical management. The treatment center becomes a temporary society and identity; substitution of a network can occur in an attempt to resolve the dilemma of loss in this area (Heine, Proulx & Vohs, 2006). The experience when a fellow patient dies promotes survivors’ guilt and renews fear of one’s mortality. Isolation increases when visits to the center are reduced, but the patient is still restricted from activities. Endurance and easy fatigability limits activity. Patients experience changes in endocrine function that affect perceived wellness. Cognitive effects caused by treatment (colloquially known as “chemo brain”) causes difficulties in function and return to work. Patients often express a fear of death; assessment of clinical depression is confounded by chronic illness, medicinal effects and existential/spiritual distress (Williams-Lloyd, Reeve, Kissane, 2008; Kissane et al., 2004). Existential distress is defined as difficulty coping accompanied by feelings of helplessness, lack of meaning, isolation, fear of mortality, reduced self-efficacy and self-esteem, and diminished hopefulness. This existential crisis can be induced by the SCT process and its resultant functional restrictions and outcomes (Morstyn, 2009; Xuereb & Dunlop, 2003).

Allogeneic transplant has the added difficulty of integrating the donor’s immune system with the host, in the hope that a graft (the donor cells) versus disease effect will occur. Transplant may be unsuccessful and fail to achieve a cure or extended disease-free survival due to relapse of underlying disease. These risks are unpredictable so maintaining a steady state which is essential for reduction in threat to meaning is not possible. This instability, combined with effects on a patient’s physical and cognitive abilities, creates challenges in engaging and sustaining meaning making attempts. Victor Frankl (1959) put forth the idea that meaning in life was central to healthy functioning and adaptation.

Patients who are transplant candidates have high risk diseases that are unlikely to remain in remission with standard therapy. Transplant with its risks is a choice made by patients hoping for cure or extended life. Clinical observation of the authors suggests that stem cell transplant threatens individual meaning (self-esteem, goals, mortality), disrupt relational networks (friends, work, community activities), and cause role loss because of treatment. The individual’s interpretation of suffering varies by culture; cultural archetypes, such as “rugged individualism,” are common in the United States, which may worsen the impact of these disruptions on a personal level (May 1991).

## METHODS

### PATIENTS

Inclusion criteria included SCT patients’ ≥ 18 years of age. Potential subjects who were developmentally or cognitively impaired were excluded. Potential subjects unable to complete the assessment instruments in English were excluded, as all instruments were written in English. Participants were offered enrolment during their pre-transplant psychosocial evaluation and were given their first set of instruments. The number of patients that met study criteria and were transplanted from March 8, 2013-March 8, 2015 totaled approximately 254; all eligible patients were approached by social workers and 146 were enrolled. The reason for non-enrolment was patients declining to participate in the study. Nine patients who were enrolled did not medically advance to the transplant process. The primary cause of patient’s dropping out of the study was death or severe morbidity preventing further completion of the survey. 32 patients died within a year or less of their transplant. Seven patients withdrew due to changing their minds about participating (they felt overwhelmed by their treatment or did not want to bother with the questionnaires). One patient was withdrawn due to not completing the first measure prior to transplant. The principal investigator was responsible for follow up to provide subsequent survey packets and reminder letters if instruments were not returned. The patients were given packets in person or through the mail based on their schedule of visits to the center. The instruments following the pre-transplant time period were provided based on their date of transplant (that determined the one, three, six month and one-year time frames) with a window of +-15 days. Questionnaires returned were entered into the data base built in REDCap (Research Electronic Data Capture Consortium) primarily by the principal investigator with assistance from a MSW graduate student (the data was de-identified at that time).

### STUDY MEASURES

Demographic data was collected by the principal investigator from the electronic medical record (Cerner) and from CIBMTR (*Center for International Blood and Marrow Transplant Research)* records kept by the program. The time periods selected for examination (baseline, one month, three months, six months, and one year) were selected in consultation with the physician advisor as clinically significant in the recovery trajectory from transplant. Response rates were calculated for each time period. This study was approved by the institution’s IRB (study number HM14968) and the Massey Cancer Center’s Protocol Review and Monitoring Committee. The patients provided written informed consent and signed HIPPA statements as required by the IRB.

The following questionnaire was used at all time points. The instrument is available as supplementary material.

The Illness Perception Questionnaire-Revised (IPQ-R; Moss-Morris et al., 2002) is a 7 domain Likert-type item questionnaire that measures various aspects of understanding of one’s illness and served as the primary measure of situational appraisal. In addition to the domains of Identity, Timeline (Timeline-acute/chronic and Timeline cyclical), Consequences, Cure/Controllability, and the Causes subscales found in the original version, the revised version includes Emotional Representation and Illness Coherence subscales which consider the cognitive and emotional aspects of illness perception (Abubakari et al., 2012). This instrument is based on Leventhal’s Self-Regulatory Model and was chosen due to its use of health-related constructs as a measure of situational appraisal (Moss-Morris et al., 2002). This is related to the meaning making model as it was used in this study to assess situational appraisal of SCT patients as this appraisal drives the experience of distress that theoretically leads to meaning making attempts (see Figure 1 for model diagram). The sub-scales are used exclusively in scoring and there is no total score for the instrument as a whole.

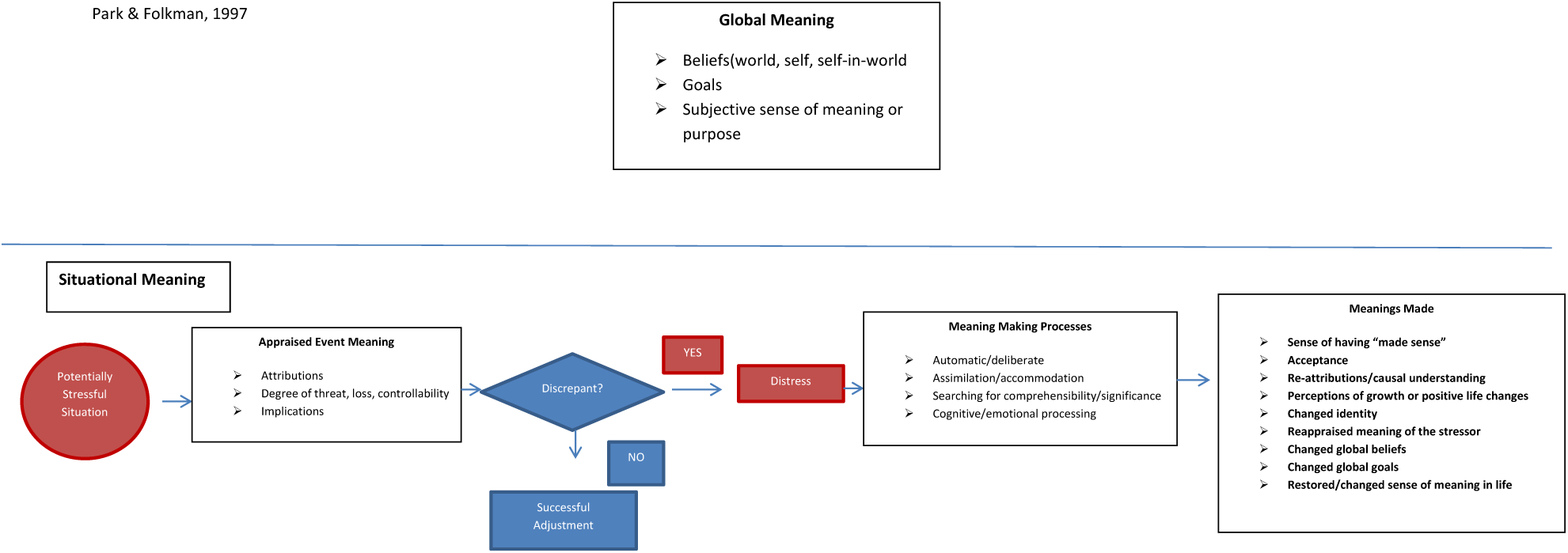

#### Subscales of the IPQ-R

The IPQ-R subscales can be sub-divided into two broad categories: those representing positive situational attribution, and those representing negative situational attribution. There are three subscales representing positive situational attribution: Treatment Control, Personal Control, and Illness Coherence. Treatment and personal control reflect the positive illness belief that one has control over treatment and one’s personal life; i.e., Examples are “Treatment can control my illness;” and “What I do can determine whether my illness gets better or worse.” Illness coherence reflects the positive illness belief that the condition makes sense; i.e., “I don’t understand my illness,” which is a personal understanding of the illness or a meta-cognition (Moss-Morris et al., 2002). Higher values for each of these three subscales indicate more positive situational attribution.

There are five subscales representing negative situational attribution: Identity, Timeline (Acute or Chronic), Timeline Cyclical, Emotional Representation, and Consequences. The Identity subscale reflects the presence of physical symptoms and whether the patient attributes them to the illness or having been present prior to their illness, including nausea, fatigue, weakness and other symptoms that are related to cancer treatment. This sub-scale separates somatization from illness identity (Moss-Morris et al., 2002). Greater illness related symptom burden relates to patients’ reporting reduced control and greater negative illness attributions. Timeline reflects the chronic nature of the illness (i.e., “my illness will last a long time”) while timeline cyclical reflects a person’s sense that their symptoms come and go and are unpredictable (i.e., “The symptoms of my illness change from day to day”). Emotional representation is the assessment of negative emotions related to the illness; i.e., “Having this illness makes me anxious.” This subscale was added to the revised version of the instrument to capture the emotional aspects of illness in addition to the cognitive aspects that are a part of the other sub-scales. Consequences reflects the assessment of negative results of an illness on one’s life; i.e., “My illness does not have much effect on my life.” Higher scores on these sub-scales reflect greater negative view of the illness.

### STATISTICAL METHODS

IPQ-R subscales for positive situational attributions (treatment and personal control, illness coherence) and negative situational attributions (timeline acute or chronic, timeline cyclical, emotional representations, and consequences) are summarized at each time point with sample sizes, means, and standard deviations. Trends in these means over time are tested using a linear mixed effect model with the IPQ-R subscale as a continuous, normally distributed outcome and a five-level fixed effect for time. Dependence within subjects are fit with a first-order auto regressive correlation structure (heterogeneous variance was examined but based on small-sample adjusted AICs did not fit as well as assuming homogeneous variance). The MEANS and GLIMMIX procedures in the SAS statistical software (version 9.4, Cary, NC, USA) were used for analysis.

#### Sample Size Determination

Assuming ten total comparisons for each IPQ-R subscale (baseline to 1 month, baseline to 3 months, baseline to 6 months, baseline to one year, 1 to 3 months, 1 to 6 months, 1 month to 1 year, 3 to 6 months, 3 months to one year, 6 months to one year), a step-down-adjusted significance level of α/k = 0.05/10 = 0.005, and a subscale standard deviation of 5.0, we planned for 136 respondents, which would provide at least 80% power for finding at least one difference of magnitude 1.6 in IPQ-R subscale means over time.

## RESULTS

### Demographics *(Table 1 here)*

Out of 148 participants (out of approximately 210 eligible patients), more patients received autologous transplants (60%) than received allogeneic transplants (40%). Most of the patients (73%) were Caucasian, while 22% were African American, which reflects the racial distribution in Virginia as a whole (Pew Research Center, 2014). There were more male participants (61%) than female (39%). In the area of educational level, 41.9% of participants had a high school education or less; 43.9% had some college. Most patients had myeloma (40%), acute leukemia (26%) or lymphoma (21%), and the others made up other blood cancer or myeloproliferative/myelodysplastic and bone marrow failure conditions. The religious background included mostly participants Protestant faiths (73%), with smaller numbers of Catholics (17%) and those with no religious tradition (8.3%). Sample sizes in Table 1 tables are at times less than 148 due to subjects missing values for particular characteristics.

**Table 1:** Patient Characteristics

| Table 1: Patient Characteristics |  |  |  |  |  |
| --- | --- | --- | --- | --- | --- |
| Sex | Frequency | Percentage | Education | Frequency | Percentage |
| Female | 51 | 38.9% | High School or Less | 62 | 41.9% |
| Male | 80 | 61.1% | At Least Some College | 65 | 43.9% |
| Total | 131 |  | At Least Some Graduate School | 21 | 14.2% |
|  |  |  | Total | 148 |  |
| Religion | Frequency | Percentage | Ethnicity | Frequency | Percentage |
| Catholic | 22 | 16.7% | Asian / Pacific Islander | 2 | 1.5% |
| Jewish | 2 | 1.5% | African American | 29 | 22.0% |
| None | 11 | 8.3% | Hispanic | 3 | 2.3% |
| Other | 1 | 0.8% | Mixed | 1 | 0.8% |
| Protestant | 96 | 72.7% | Other | 1 | 0.8% |
| Total | 132 |  | Caucasian | 96 | 72.7% |
|  |  |  | Total | 132 |  |
| Diagnosis | Frequency | Percentage | Donor Type | Frequency | Percentage |
| AA | 1 | 0.8% | Allo | 51 | 40.5% |
| ALL | 4 | 3.0% | Auto | 75 | 59.5% |
| AML | 20 | 15.2% | Total | 126 |  |
| CLL | 4 | 3.0% |  |  |  |
| CML | 6 | 4.6% | Type of Preparation | Frequency | Percentage |
| HD | 6 | 4.6% | Full Intensity | 105 | 80.2% |
| MDS | 8 | 6.1% | Reduced Intensity | 26 | 19.8% |
| MF | 3 | 2.3% | Total | 131 |  |
| MM | 53 | 40.2% |  |  |  |
| NHL | 22 | 16.7% |  |  |  |
| Other | 5 | 3.8% |  |  |  |
| Total | 132 |  |  |  |  |

### Overall Trends in Situational Attribution

Response rates for the IPQ-R subscales ranges between 43%-93% at baseline, 26%-72% at time 2, 30%-62% at time 3, 29%-56% at time 4, 16%-45% at time 5, and 3%-5% at time 6. Longitudinal trends in IPQ-R subscales are found in Table 2. For the Positive Situational Attribution subscales, there was significant evidence of change over time in the Treatment Control and Personal Control scales. Participants’ scores trended downwards for treatment control over time (F (4,347.7) = 9.1; p=<0.0001). Personal Control also trended downwards less so than Treatment Control (F (4,336.1) =3.0; p=0.0203).The patients reported that their perception of control over their treatment decreased throughout the study year. For the Negative Situational Attribution subscales, only Consequences showed a slow but significant decrease over time (F (4,346.7) = 3.1; p=0.0166). Participants views of how much the illness had negative consequences in their lives decreased over the study year. There was non-significant evidence for changes in Timeline Acute/Chronic and for Emotional Representations. The patients viewed their illness as more chronic as time went on – scores increased over the study year (F (4,329.6) = 2.1; p=0.0835). Emotional Representations scores started high and then were lowest by 90 days, followed by an increase that ended at a year out higher than but not as high as pre-transplant measures (F (4,346.8) = 2.3; p=0.0567). The patients’ negative feelings about their illness were lowest at 90 days.

**Table 2:**
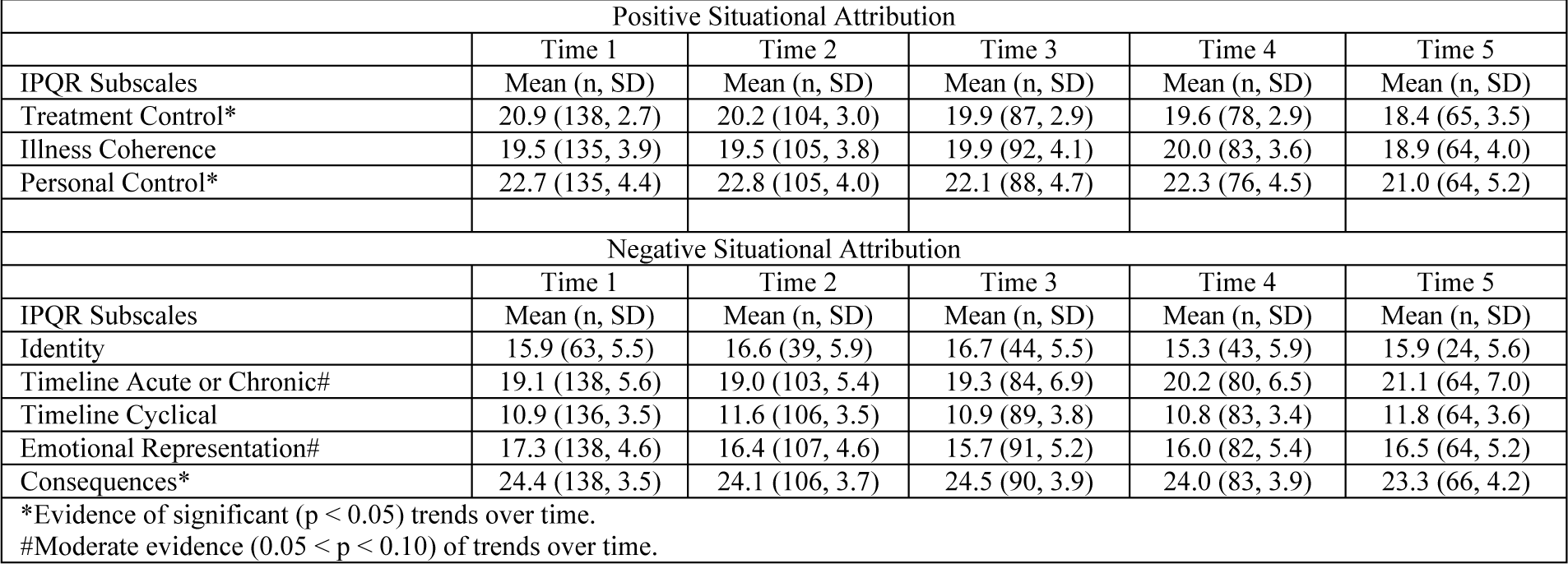
Overall trends in Situational Attribution

### Trends by Participant Characteristics

Trends in IPQ-R subscales over time are presented for different levels of patient characteristics (when significant) in Table 3. *(Donor Type)* The Treatment Control subscale did not change over time based on donor type, through there was a significant difference between groups (F (1,149.0) = 8.8; p=0.0035). Allogeneic patients reported feeling more control over their treatment than autologous patients. There were significant Timeline Cyclical trends over time for donor types (F (4,305.0) = 2.5; p=0.0411), with subscale scores occasionally increasing in allogeneic patients and remaining relatively constant in autologous patients. There was no significance in trends in the Consequences subscale over time, but there were significant differences between donor types (F (1,151.9) = 6.1; p=0.0148), where allogeneic patients had higher scores and so viewed their illness as having more consequences on their lives, then autologous patients.

**Table 3:**
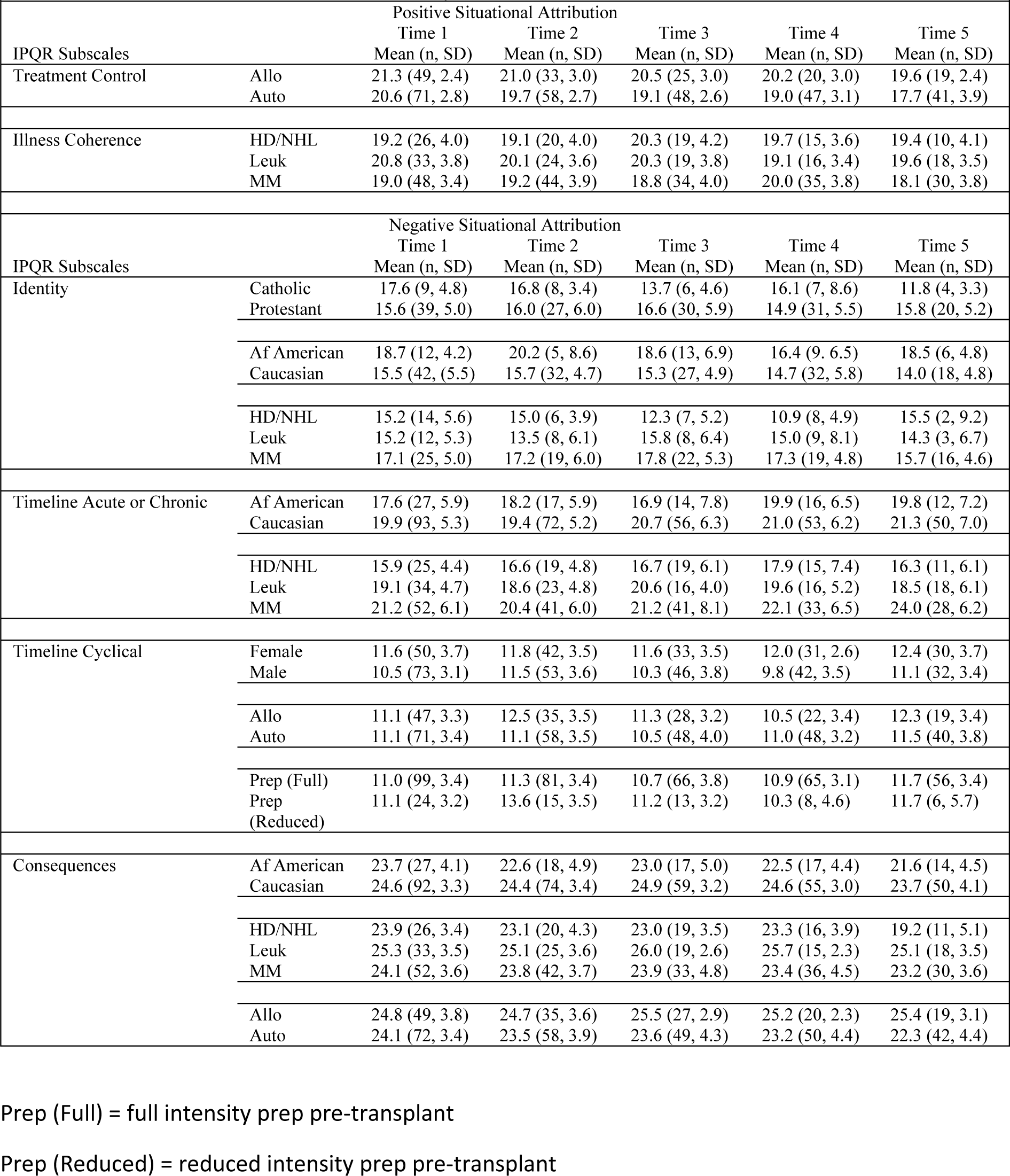
Trends in Situational Attribution by Patient Characteristics

*(Diagnosis)* There were significant differences in the Illness Coherence subscale over time for diagnostic groups (F (8,281.6) = 2.3; p=0.0203). The lymphoma patients’ scores stayed within the same range while leukemia patients’ scores decreased, and myeloma patients showed a one-time increase at 180 days. Leukemia patients felt that their illness made less sense then lymphoma and myeloma patients. There were no significant trends in the Timeline subscale over time for diagnostic groups, though there was a significant difference between groups on this measure (F (2,136.7) = 6.7; p=0.0016). Myeloma patients viewed their illness as more chronic than the other groups. Leukemia patients were the next highest group, with lymphoma patients viewing their illness as less chronic and more acute. Likewise, there was no significant trends in the Consequences subscale over time for diagnostic groups and attribution of consequences, though there were differences between the groups (F (2,134.0) = 5.7; p=0.0041). Leukemia patients viewed the illness as having the most consequences on their lives, myeloma patients were next highest, followed by lymphoma patients.

*(Ethnicity)* There were significant differences in the Timeline subscale over time between Caucasian and African American patients in how they viewed the illness chronicity (F (4,285.4) = 3.9; p=0.0041). African Americans began with a lower score which increases over time – they saw their illness as more acute in the beginning and more chronic later. Caucasians viewed their illness as a more chronic phenomenon early on and this increased at a slower pace than their African American counterparts. All scores trended to indicate they viewed their illness as more chronic as the study year progressed. There was no difference in temporal trend in the Consequences subscale between ethnic groups, but there were absolute differences in the scores between groups (F (1,136.4) = 5.5; p=0.0211). Caucasian patients viewed their illness as having greater consequences for their lives while African Americans viewed the illness has having less of an impact.

*(Gender)* There were significant differences in the Timeline Cyclical subscale between men and women’s views of the illness as coming and going and being unpredictable (F (1,143.9) = 6.7; p=0.0106). Women viewed the cyclical and unpredictable nature of the illness as greater than men.

*(Type of Preparation (treatment prior to cell infusion))* There were significant differences in the Timeline Cyclical subscale over time for patients who had the two types of pre-transplant preparation regimens (full intensity vs. reduced intensity) (F (4,323.0) = 2.7; p=0.0332). The reduced intensity group included those who did not have fully myeloablative transplants. The patients with reduced intensity transplants had higher scores on this measure at day 30 – they viewed their illness as unpredictable and changeable – then those with full intensity treatment.

## DISCUSSION

This study showed change over time in measures that illustrated the SCT experience. The focus was on situational attribution. Park and George (2013) speak to the challenges of examining the complex and theoretically rich concepts inferred in the meaning making model. Change over time was found within the group as a whole, donor types, different diagnoses, within the two major ethnic groups, between genders, and those patients with different pre-transplant prep regimens.

The meaning making model posits that if meaning is made, then distress is relieved and the cycle of meaning making attempts can be resolved. The nature of stem cell transplant – especially for those that receive donor cells – lends itself to a long process and unexpected complications. Patients and their families often remark that it is difficult to make short or long-term plans. Park and Gutierrez (2013) discussed that well-being was strongly associated with one’s perception of controllability of circumstances. In our study, participants’ felt that treatment control decreased over time. Appraisal violations of global meaning by attribution of a situation as traumatic lead to stress; emotional well-being was worse pre-transplant as patients cope with shock, the unknown, and feel anxiety about the upcoming transplant (Park, Mills & Edmondson, 2012). Cure of disease does not mean life lacking disability or residual effect - patients viewed their illness as more chronic as time passed. This indicates the varied wellness and experience of the patients as frequent meaning violations create greater physiologic and psychological stress (Park, Mills & Edmondson, 2012; Steger, 2012).

The effects of treatment protocol explain the variance seen between diagnostic groups. African American patients attributed less effect of the illness on their lives than Caucasian patients (consequences) but attributed more of the symptoms to their illness (higher Identity scores). The numbers of African Americans in the study who completed the Identity scores was low, limiting the conclusions drawn. They also started out seeing their illness as less chronic.

Women saw their illness as unpredictable compared to men. The reason for this is unclear and the study itself did not answer this question. Socialized gender differences and physiology could be explanations.

Patients who received a less intense preparative regimen perceived their illness to be unpredictable and changing. This appears counterintuitive as one would assume a less medically toxic protocol would infer fewer side effects from toxicity, hence more predictable and less changeable symptoms.

### Other Results - Discussion

There were patient characteristics that lacked change over time and did not have absolute differences in IPQ-R sub-scales. If differences based on a characteristic were stable over time, this may be indicative of resiliency factors. Hayes (Hayes, Laurenceau, Feldman, Strauss & Cardaciott, 2007) and Bonanno (2004) suggest, there are non-linear processes at work and intermittent distress can occur, but psychologically healthy individuals remain stable over time. SCT pre-transplant evaluation screens out patients psychosocially and medically not equipped to manage the process as ethics dictates one not cause harm. Hayes argues for single subject design as a population contains individuals with differing recovery trajectories (Hayes, Laurenceau, Feldman, Strauss & Cardaciott, 2007). The results above argue for study of discrete populations, determining which patients have strong resilience characteristics, and how this affects their process. Meaning making may not always occur. The authors’ experience clinically gives witness to individuals’ non-linear experience and sudden change in coping (both in onset of distress and in post-traumatic growth). The group analysis masked the individualized experience.

### Study Limitations

The study of meaning making is theoretically rich and methodologically challenging; some of the challenges could not be controlled adequately in this study (Park, 2010). Participants’ responses are related to a variety of factors – whether they were responding to their cancer not the transplant, possibility of cure, and specific complications vs. the transplant experience. These are confounding factors. Not all patients completed the entire set of questionnaire packets. Patient mortality affected this result - occasionally patients were too sick at intervals to complete instruments. There are differences between autologous and allogeneic patients or those with malignancies, and those who had bone marrow failure/other non-malignant conditions (Braamse, Gerrits, Meijl, 2012). The pre-transplant course of treatment and disease characteristics are different in some cases. Study of discrete groups within the transplant population reduces sample size thus reducing statistical significance. Multi-center studies are indicated here.

The questionnaire packets were self-administered and not done in the presence of the investigators. Enrollment was conducted by the social workers on the study who handled the distribution of study instruments in person or through the mail. The combination of serving as clinical treatment staff and investigating staff could have introduced response bias; there is no indication that this occurred. The conditions under which participants completed questionnaires varied over their study year (in hospital, clinic, or home). Some patient groups had small numbers - the tandem transplant patients and those receiving reduced intensity conditioning. Analyses of the smaller numbered subsets limit data generalizations. Occasionally participants complained about the repetition of measures over time.

This study involved combining behavioral health measures with medical data. The realities and structure of data management at the Health System prevented the researchers from gathering specific medical data that might have better clarified results. Data for SCT patients was maintained in more than one system. These systems were not connected; the type and quality of data available varied. The data was recorded by more than one provider and was sometimes incomplete.

### Clinical Implications

Clinical observation indicates the recovery from transplant does not occur in a linear fashion and is marked by complications that arise unexpectedly. This suggests that the participants may re-experience distress due to the repeated crises affecting the ability to maintain a steady state needed to accommodate or assimilate meaning (Davis, Wortman, Lehman & Cohen-Silver, 2000; Park, 2010). This is due to perception of threat vs. controllability in coping to manage the challenge (Bonanno, 2013). There is evidence that patient’s recovery is affected by ability to participate in tasks, like exercise (Fiuza-Luces, Simpson, Ramirez, Lucia & Berger, 2016) and this can be driven by expectation set and coping. Continued threat leads to continued distress (Park & Al, 2006). Significant demographic differences lacking change over time infer stable characteristics that argue for study of resilience factors. This study indicated ethnic differences in illness attribution which are under examined in the literature. What are affects that exist in social oppression and inequity? How do social determinants of health and resilience factors affect the patients? Variability occurs as a result of culture, age, gender and spirituality (Alea & Bluck, 2013; Park, 2005; Hart & Singh, 2009). There is evidence that there are differing predictors and correlates these include, but are not limited to: meaning, purpose, well-being, accommodation, assimilation, post-traumatic growth and post-traumatic depreciation (Kissane, et al., 2004). Future studies of situational attribution, resilience, and regulatory flexibility might be indicated.

Goals and beliefs are postulated to make up one’s global meaning. Violations of global meaning by situational appraisal involve conscious and automatic meaning making attempts (Xuereb & Dunlop, 2003). Researchers can examine meanings made and mechanisms that facilitate this end (accommodation vs. assimilation). Global meaning and post-traumatic growth were not directly studied by these authors. It would be helpful to assess the type of meanings made by SCT patients and how this effects adjustment. Inclusion of measures that determine incidents of repeated crises in the patients’ medical trajectory is recommended. Patients that derive a sense of purpose, reason and meaning to the suffering they experience and that have renewed relationships and post-traumatic growth that incorporate the challenges of cancer are important for successful adjustment (Park, 2010; Davis, Wortman, Lehman & Cohen-Silver, 2000). Combining resilience work and meaning making has application here. These are areas worth future research, and SCT patients enable the option of prospective and longitudinal study.

## Data Availability

data housed in Red Cap via Vanderbilt

---

Prep (Full) = full intensity prep pre-transplant Prep (Reduced) = reduced intensity prep pre-transplant
| Survey Completion Rates over time |  |  |  |
| --- | --- | --- | --- |
| Time | IPQ-Identify | IPQ-Timeline | IPQ-Consequences |
| 1 | 63 (43%) | 138 (93%) | 138 (93%) |
| 2 | 39 (26%) | 103 (70%) | 106 (72%) |
| 3 | 44 (30%) | 84 (57%) | 90 (61%) |
| 4 | 43 (29%) | 80 (54%) | 83 (56%) |
| 5 | 24 (16%) | 64 (43%) | 66 (45%) |

| Survey Completion Rates over time |  |  |  |  |  |
| --- | --- | --- | --- | --- | --- |
| Time | IPQ-Personal Control | IPQ-Treatment Control | IPQ-Illness Coherence | IPQ-Timeline Cyclical | IPQ-Emotional Representation |
| 1 | 135 (91%) | 138 (93%) | 135 (91%) | 136 (92%) | 138 (93%) |
| 2 | 105 (71%) | 104 (70%) | 105 (71%) | 106 (72%) | 107 (72%) |
| 3 | 88 (59%) | 87 (59%) | 92 (62%) | 89 (60%) | 91 (61%) |
| 4 | 76 (51%) | 78 (53%) | 83 (56%) | 83 (56%) | 82 (55%) |
| 5 | 64 (43%) | 65 (44%) | 64 (43%) | 64 (43%) | 64 (43%) |

